# Prevalence of nutritional, behavioral and anthropometric cancer-related risk factors among adults in Nouakchott, Mauritania: a cross-sectional study

**DOI:** 10.64898/2026.05.23.26353924

**Authors:** Nah Tolba, Adil Najdi, Mohamed El Hfid, Mohameddou H’meied Maham, Selma Mohmed Brahim, Ahmedou Tolba, Nabila Sellal

## Abstract

**Background:** Cancer is a growing public health challenge in low- and middle-income countries, where urbanization, nutritional transition and lifestyle changes contribute to modifiable risk factors. In Mauritania, population-based data on cancer-related nutritional, behavioral and anthropometric risk factors remain limited.

**Objective:** To describe the frequency of the main nutritional, behavioral and anthropometric cancer-related risk factors among adults living in the three wilayas of Nouakchott.

**Methods:** A cross-sectional study was conducted among 1,000 adults aged 18 years and older in Nouakchott. Data were collected using a standardized questionnaire covering sociodemographic characteristics, dietary habits, physical activity and selected health behaviors. Anthropometric measurements were performed to assess body mass index and abdominal adiposity. Abdominal obesity was defined using sex-specific waist circumference cut-off points recommended by the World Health Organization: ≥ 88 cm in women and ≥ 102 cm in men. Results were presented as frequencies and proportions, with comparisons by sex, age group and wilaya of residence.

**Results:** Women represented 52.0% of participants, and 53.5% were aged 18–34 years. Excess body weight was frequent, with 38.6% overweight and 28.0% obese. Abdominal adiposity was also common, with 58.0% having increased or substantially increased waist circumference and 48.3% having an elevated waist-to-hip ratio. Physical inactivity was reported by 64.7% of participants, and 15.7% were current smokers. Dietary exposures included high red meat consumption in 66.8%, daily refined cereal intake in 67.5%, daily sugar-sweetened beverage consumption in 14.9%, and limited daily fresh fruit consumption in 13.8%. Significant differences were observed by sex for anthropometric indicators, by age for selected dietary habits, and by wilaya for physical activity, smoking and selected dietary behaviors.

**Conclusion:** This study shows a high frequency of modifiable cancer-related risk factors among adults in Nouakchott, particularly excess body weight, abdominal adiposity, physical inactivity and unfavorable dietary habits. These findings support the need to strengthen primary prevention strategies targeting nutrition, physical activity and tobacco control in Mauritania.

## 1 Introduction

Cancer is a major global public health problem because of its growing burden on morbidity, mortality, and health systems. In 2020, it was responsible for nearly 10 million deaths worldwide, with a particularly heavy burden in low- and middle-income countries, where approximately 70% of cancer-related deaths occur [1]. Epidemiological data show that a substantial proportion of cancers is linked to modifiable risk factors, particularly behavioral, nutritional, and environmental factors. Smoking, unhealthy dietary habits, physical inactivity, and obesity are now recognized as important risk factors for several cancer sites [2].

Over recent decades, urbanization, the globalization of consumption patterns, and sociocultural changes have profoundly altered population lifestyles. These changes are reflected in increased consumption of processed foods high in sugar, salt, and fat, reduced physical activity, and a rise in overweight and obesity. In this context, the relationship between diet, physical activity, body weight, and cancer risk has become central to prevention strategies [2,3].

Mauritania is no exception to this nutritional and epidemiological transition. As in many low-and middle-income countries, the rise in noncommunicable diseases is driven by rapid urbanization, changing lifestyles, and shifts in dietary patterns. In 2022, the country recorded 3,274 new cancer cases and 2,234 deaths attributable to this disease [4]. This situation is particularly concerning because available data on cancer risk factors in Mauritania remain limited, fragmented, and insufficiently documented, although several common cancers in the country may be influenced by modifiable factors.

In this context, it is essential to better document nutritional and behavioral cancer-related risk factors in the Mauritanian population. The present study, conducted in the three wilayas of Nouakchott, aimed to describe the frequency and distribution of the main nutritional and behavioral risk factors associated with cancer in the general population.

This cross-sectional study constitutes the descriptive component of a broader doctoral research program on nutritional, behavioral and anthropometric cancer-related risk factors in Nouakchott. A complementary case-control study was conducted simultaneously to identify factors independently associated with colorectal cancer in the same urban setting.

## 2 Methods

### 2.1 Study design and setting

This was a cross-sectional study conducted in the three wilayas of Nouakchott: Nouakchott North, Nouakchott South, and Nouakchott West.

### 2.2 Study period

The survey was conducted from January 25 to April 25, 2026, in the three wilayas of Nouakchott.

### 2.3 Study population and selection criteria

The study population consisted of Mauritanian adults aged 18 years and older who were residing in the three wilayas of Nouakchott at the time of the survey.

All Mauritanian adults aged 18 years or older, residing in Nouakchott, who provided informed consent to participate were included in the study.

Individuals under 18 years of age, pregnant women, bedridden persons, and individuals with mental disorders preventing them from adequately answering the questionnaire were not included.

### 2.4 Sampling

#### 2.4.1 Sample size

The sample size was estimated using the standard formula for prevalence studies:

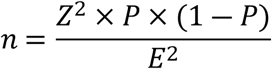

In the absence of recent and specific population-based data on the prevalence of the main nutritional and behavioral cancer-related risk factors in Nouakchott, an expected prevalence of 50% was used. This choice, commonly applied when the expected prevalence is unknown, maximizes the required sample size and provides a conservative estimate of the necessary number of participants.

The calculation was based on a 95% confidence level (Z = 1.96), an absolute precision of 4%, and an expected prevalence of 50%. The initial required sample size was therefore estimated at approximately 600 participants. To account for the cluster sampling design, a design effect of 1.5 was applied, increasing the minimum required sample size to approximately 900 participants. Considering the operational constraints of fieldwork, the need to ensure adequate coverage of the three wilayas of Nouakchott, and the need for a sufficient sample size for descriptive and stratified analyses, the final sample size was increased to 1,000 participants. Participants were distributed across 50 clusters of 20 individuals.

#### 2.4.2 Sampling procedures

Stratification was performed according to the three wilayas of Nouakchott: Nouakchott North, Nouakchott West, and Nouakchott South, each constituting a separate stratum.

In the first stage, the moughataas were considered the primary sampling units. Nouakchott comprises nine moughataas distributed across the three wilayas. Clusters were selected in proportion to the population size of each wilaya, in order to ensure balanced coverage of the capital.

In the second stage, households were randomly selected within the selected clusters, from accessible housing units in the survey areas.

In each selected household, only one participant aged 18 years or older was included. When several household members were eligible, the participant was selected randomly according to a standardized procedure.

Before data collection began, a preparatory phase was conducted to test the tools, identify potential logistical constraints, and harmonize procedures. The interviewers were then trained on the sampling method, questionnaire administration, and the practical procedures for data collection.

### 2.5 Data collection tools and techniques

Data were collected using a structured questionnaire specifically developed for this study. This tool was used to collect relevant sociodemographic, nutritional, behavioral, and anthropometric information from participants who met the inclusion criteria.

The questionnaire was digitized and administered through the KoboCollect application, allowing direct, standardized, and secure data entry in the field. The use of this tool helped improve data quality, reduce data entry errors, and facilitate data centralization.

Data were collected prospectively by previously trained interviewers using mobile devices equipped with KoboCollect. At the end of the fieldwork, the data were exported to Excel format for verification, cleaning, and preparation for statistical analysis.

### 2.6 Data management and analysis

After data collection, quality control procedures were performed, including the identification and removal of duplicates, detection of outliers and verification against available sources, as well as variable coding for analysis. High consumption of dietary items was defined as consumption at least three times per week, corresponding to the categories “3–4 times/week” and daily. The data were then processed and analyzed using Epi Info™ version 7.2.4.0 and IBM SPSS Statistics, version 22 (IBM Corp., Armonk, NY, USA), while tables and graphs were prepared using Microsoft Excel 365.

A descriptive analysis was conducted to summarize the sociodemographic, nutritional, behavioral and anthropometric characteristics of the study population. Qualitative variables were presented as frequencies and percentages. Subsequently, stratified analyses were performed to examine the distribution of the main risk factors according to selected sociodemographic variables, particularly sex, age group and wilaya of residence. Comparisons between qualitative variables were performed using Pearson’s chi-square test. Statistical significance was set at p < 0.05. No substantial missing data were observed for the variables included in the final analysis.

### 2.7 Ethical considerations

This study was conducted in accordance with the ethical principles applicable to research involving human participants, in line with the Declaration of Helsinki. The research protocol received a favorable opinion from the Health Research Ethics Committee of the Ministry of Health of Mauritania, under reference 047-2026/MS/CNERS, dated January 22, 2026.

Participation in the study was free and voluntary. Before inclusion, all participants were informed about the objectives of the research, the study procedures, and how the collected data would be used. Informed consent was obtained from each participant. Participants were free to refuse participation or withdraw from the study at any time, without any consequences.

Confidentiality and anonymity were ensured throughout the study. The collected data were coded, without any direct personal identifiers, and used exclusively for scientific purposes.

## 3 Results

### 3.1 General characteristics of participants

A total of 1,000 adults from the three wilayas of Nouakchott were included in the study. Women represented 52.0% of the sample, and the study population was relatively young, with 53.5% of participants aged 18–34 years.

Most participants resided in Nouakchott South (43.0%) or Nouakchott North (42.0%), while Nouakchott West accounted for 15.0% of the sample. The population had a relatively high educational level, with 61.2% reporting higher education. However, unemployment was frequent, affecting 41.2% of participants.

Anthropometric indicators showed a high burden of excess body weight and abdominal adiposity. Overall, 66.6% of participants were overweight or obese, 58.0% had an increased or substantially increased waist circumference, and 48.3% had an elevated waist-to-hip ratio.

Overall, the sample was largely young and highly educated, but showed a high prevalence of overweight, obesity and abdominal adiposity, suggesting a substantial burden of metabolic and cancer-related risk factors in Nouakchott. Other general characteristics are presented in Table 1.

**Table 1.** General characteristics of participants (N = 1,000)

| **Variable** | **Category** | **Frequency (n)** | **Percentage (%)** |
| --- | --- | --- | --- |
| **Sex** | Male | 480 | 48.0 |
|  | Female | 520 | 52.0 |
| **Age group, years** | 18–24 | 249 | 24.9 |
|  | 25–34 | 286 | 28.6 |
|  | 35–44 | 178 | 17.8 |
|  | 45–54 | 132 | 13.2 |
|  | 55–64 | 90 | 9.0 |
|  | ≥65 | 65 | 6.5 |
| **Wilaya of residence** | Nouakchott North | 420 | 42.0 |
|  | Nouakchott West | 150 | 15.0 |
|  | Nouakchott South | 430 | 43.0 |
| **Education level** | No formal education | 142 | 14.2 |
|  | Primary | 83 | 8.3 |
|  | Secondary | 163 | 16.3 |
|  | Higher education | 612 | 61.2 |
| **Marital status** | Single | 622 | 62.2 |
|  | Married | 292 | 29.2 |
|  | Divorced/widowed | 86 | 8.6 |
| **Occupation** | Unemployed | 412 | 41.2 |
|  | Student | 119 | 11.9 |
|  | Trader/artisan | 116 | 11.6 |
|  | Employee/civil servant | 277 | 27.7 |
|  | Other | 76 | 7.6 |
| **Body mass index** | Underweight | 54 | 5.4 |
|  | Normal weight | 280 | 28.0 |
|  | Overweight | 386 | 38.6 |
|  | Obesity | 280 | 28.0 |
| **Waist circumference** | Normal | 420 | 42.0 |
|  | Increased risk | 297 | 29.7 |
|  | Substantially increased risk | 283 | 28.3 |
| **Waist-to-hip ratio** | Normal | 517 | 51.7 |
|  | Elevated | 483 | 48.3 |

### 3.2 Main nutritional and behavioral exposures among participants

Several potentially unfavorable dietary and behavioral exposures were common in the study population. Red meat was consumed at least three times per week by 66.8% of participants, while processed meat was consumed at this frequency by 15.1%. Daily consumption was particularly frequent for refined cereals (67.5%), followed by foods rich in refined sugars (39.9%), salty foods (37.9%), canned foods (16.8%), sugar-sweetened beverages (14.9%) and fried foods (14.2%).

The consumption of foods associated with cancer risk reduction was variable. Daily intake was reported by 43.4% of participants for fresh vegetables, 43.7% for dairy products, 14.4% for fish and only 13.8% for fresh fruit. Conversely, 20.9% of participants reported never consuming fresh fruit, and 23.4% reported never consuming fresh vegetables.

Regarding behavioral factors, current smoking was reported by 15.7% of participants, whereas no participant reported alcohol consumption. Physical activity was generally low, with 64.7% reporting no regular physical activity. Walking was the most commonly reported activity, but concerned only 22.8% of participants. Sun exposure was frequent, with only 1.6% reporting no daily exposure; however, the most commonly reported duration was less than 30 minutes per day (39.1%). Detailed results are presented in Table 2.

**Table 2.** Main nutritional and behavioral exposures among participants (N = 1,000)

| Variable | Category | Frequency (n) | Percentage (%) |
| --- | --- | --- | --- |
| **Red meat** | Never | 120 | 12.0 |
|  | 1–2 times/week | 212 | 21.2 |
|  | 3–4 times/week | 264 | 26.4 |
|  | Daily | 404 | 40.4 |
| **Processed meat** | Never | 619 | 61.9 |
|  | 1–2 times/week | 230 | 23.0 |
|  | 3–4 times/week | 58 | 5.8 |
|  | Daily | 93 | 9.3 |
| **Fried foods** | Never | 321 | 32.1 |
|  | 1–2 times/week | 413 | 41.3 |
|  | 3–4 times/week | 124 | 12.4 |
|  | Daily | 142 | 14.2 |
| **Foods rich in refined sugars** | Never | 316 | 31.6 |
|  | 1–2 times/week | 190 | 19.0 |
|  | 3–4 times/week | 95 | 9.5 |
|  | Daily | 399 | 39.9 |
| **Sugar-sweetened beverages** | Never | 383 | 38.3 |
|  | 1–2 times/week | 332 | 33.2 |
|  | 3–4 times/week | 136 | 13.6 |
|  | Daily | 149 | 14.9 |
| **Salty foods** | Never | 378 | 37.8 |
|  | 1–2 times/week | 177 | 17.7 |
|  | 3–4 times/week | 66 | 6.6 |
|  | Daily | 379 | 37.9 |
| **Canned foods** | Never | 432 | 43.2 |
|  | 1–2 times/week | 309 | 30.9 |
|  | 3–4 times/week | 91 | 9.1 |
|  | Daily | 168 | 16.8 |
| **Refined cereals** | Never | 132 | 13.2 |
|  | 1–2 times/week | 103 | 10.3 |
|  | 3–4 times/week | 90 | 9.0 |
|  | Daily | 675 | 67.5 |
| **Fresh fruit** | Never | 209 | 20.9 |
|  | 1–2 times/week | 525 | 52.5 |
|  | 3–4 times/week | 128 | 12.8 |
|  | Daily | 138 | 13.8 |
| **Fresh vegetables** | Never | 234 | 23.4 |
|  | 1–2 times/week | 253 | 25.3 |
|  | 3–4 times/week | 79 | 7.9 |
|  | Daily | 434 | 43.4 |
| **Fish** | Never | 154 | 15.4 |
|  | 1–2 times/week | 539 | 53.9 |
|  | 3–4 times/week | 163 | 16.3 |
|  | Daily | 144 | 14.4 |
| **Dairy products** | Never | 204 | 20.4 |
|  | 1–2 times/week | 233 | 23.3 |
|  | 3–4 times/week | 126 | 12.6 |
|  | Daily | 437 | 43.7 |
| **Current smoking**<br>**(cigarettes/day)** | No | 843 | 84.3 |
|  | <5 | 52 | 5.2 |
|  | 5–10 | 40 | 4.0 |
|  | 10–20 | 46 | 4.6 |
|  | >20 | 19 | 1.9 |
| **Alcohol consumption** | No | 1,000 | 100.0 |
|  | Yes | 0 | 0.0 |
| **Frequency of physical activity** | None | 647 | 64.7 |
|  | 1–2 times/week | 195 | 19.5 |
|  | 3–4 times/week | 112 | 11.2 |
|  | ≥5 times/week | 46 | 4.6 |
| **Type of physical activity** | None | 647 | 64.7 |
|  | Walking | 228 | 22.8 |
|  | Running | 39 | 3.9 |
|  | Gymnastics | 21 | 2.1 |
|  | Cycling | 7 | 0.7 |
|  | Other | 58 | 5.8 |
| **Daily duration of sun exposure** | None | 16 | 1.6 |
|  | <30 minutes/day | 391 | 39.1 |
|  | 30–60 minutes/day | 339 | 33.9 |
|  | 1–2 hours/day | 168 | 16.8 |
|  | >2 hours/day | 86 | 8.6 |

### 3.3 Stratified analysis by sex, age group and wilaya of residence

Stratified analysis showed significant differences in anthropometric indicators by sex. Women had a higher prevalence of obesity than men (40.0% vs. 15.0%; p < 0.001), as well as a higher prevalence of combined overweight/obesity (81.9% vs. 50.0%; p < 0.001). Increased or substantially increased waist circumference was also markedly more frequent among women (81.7% vs. 32.3%; p < 0.001). Conversely, an elevated waist-to-hip ratio was more common among men than women (67.5% vs. 36.5%; p < 0.001).

Several dietary exposures varied significantly by age group. High red meat consumption was highest among participants aged 55–64 years (75.6%; p < 0.001), while high fresh vegetable consumption was highest in the 45–54-year age group (62.9%; p = 0.030). High dairy product consumption was most frequent among participants aged 65 years or older (67.7%; p < 0.001).

Significant differences were also observed by wilaya of residence. Regular physical activity was lowest in Nouakchott West (24.0%; p = 0.007), whereas current smoking was highest in Nouakchott South (18.6%; p = 0.035). High processed meat consumption and high canned food consumption were most frequent in Nouakchott North, at 19.0% and 30.9%, respectively. High fresh vegetable consumption was highest in Nouakchott South (57.2%; p < 0.001). Detailed results are presented in Table 3.

**Table 3.**
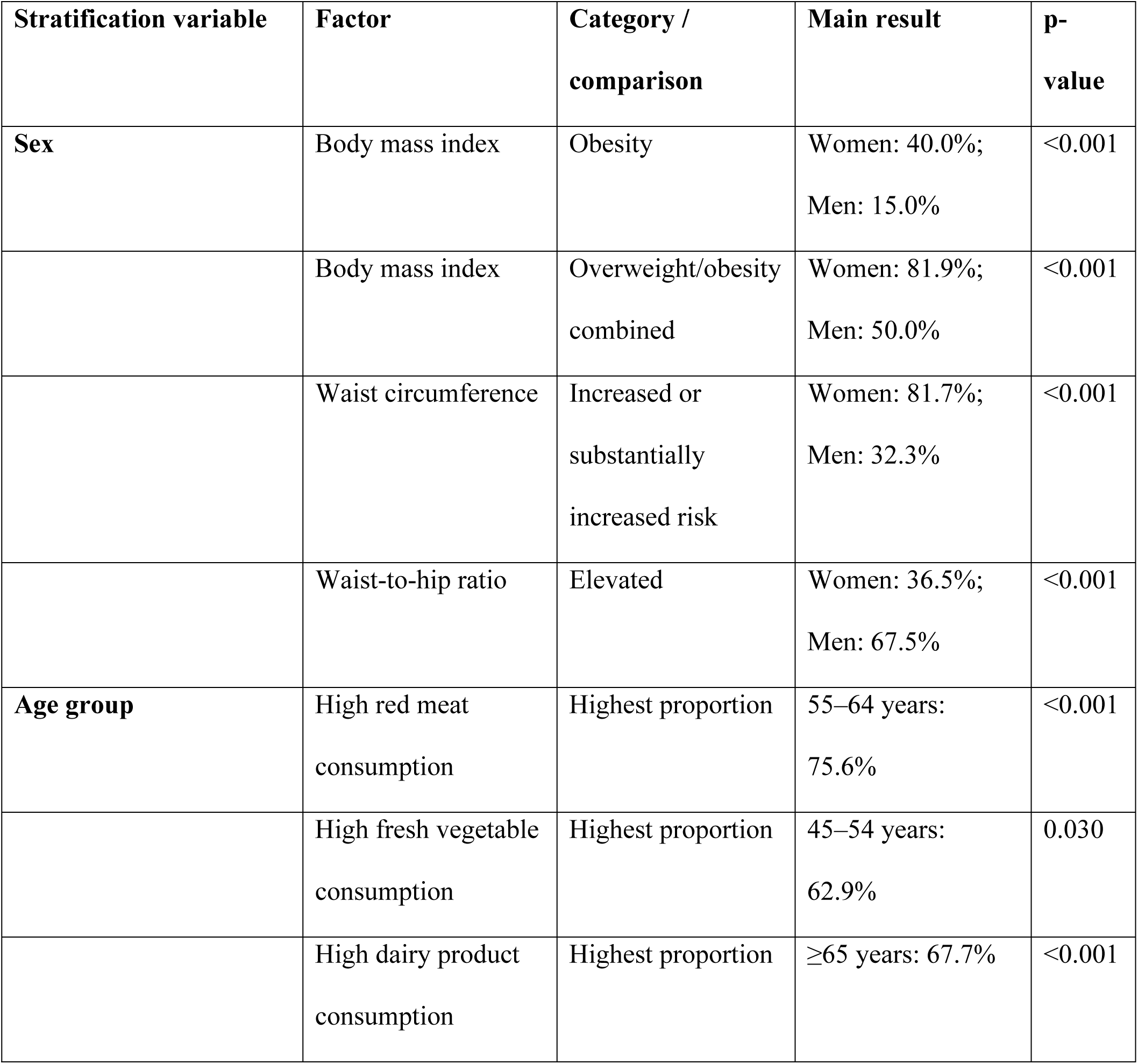
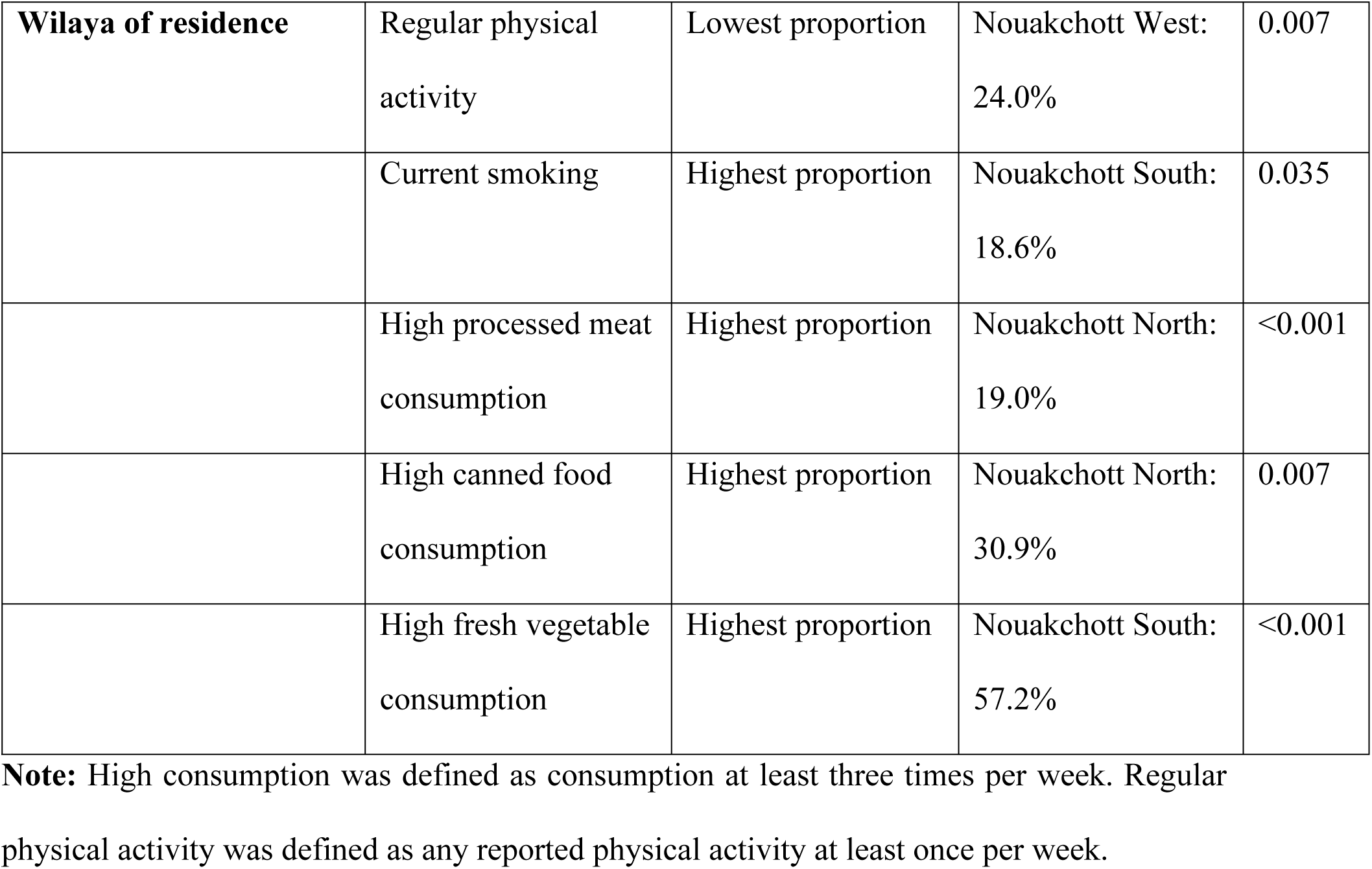
Significant stratified findings by sex, age group and wilaya of residence.

## 4 Discussion

The present cross-sectional study conducted among 1,000 adults in the three wilayas of Nouakchott highlights a high frequency of nutritional and behavioral cancer-related risk factors, despite a predominantly young study population. The main findings relate to the high frequency of excess body weight and abdominal adiposity, the low level of physical activity, and a dietary profile characterized by frequent consumption of red meat, sugar-sweetened beverages, and refined cereals, together with limited daily consumption of fresh fruit.

These findings are consistent with the nutritional transition described in many low- and middle-income countries, characterized by increasing obesity, declining physical activity, and changes in dietary habits [5,6]. This is particularly relevant given that excess body weight and certain unhealthy dietary habits are recognized as factors associated with several cancer sites, particularly colorectal, postmenopausal breast, and endometrial cancers [2,7].

The differences observed according to sex, age, and wilaya suggest that these risk factors are not homogeneously distributed within the study population. They support a differentiated interpretation of exposure profiles within the urban setting of Nouakchott.

Regarding anthropometric indicators, 66.6% of participants had excess body weight, including 38.6% who were overweight and 28.0% who were obese. This prevalence appears high compared with the global estimates reported by Afshin et al. in the GBD 2015 analysis, in which adult obesity was estimated at approximately 13% worldwide [5]. These findings are consistent with the nutritional transition observed in sub-Saharan Africa, where Steyn and McHiza reported female obesity prevalences generally ranging from 15% to 40%, with higher levels in urban settings [8].

The female predominance observed in the present study was marked: obesity affected 40.0% of women compared with 15.0% of men. This difference is consistent with regional data. In Tunisia, Ati et al. reported in 2012 an obesity prevalence of approximately 37% among women compared with 14% among men [9]. In Senegal, Macia et al. observed in 2010 an obesity prevalence of 13% among women compared with 3.9% among men in Dakar [10].

Abdominal adiposity was also frequent: 58.0% of participants had a high-risk waist circumference, including 28.3% with a very high risk, and 48.3% had an elevated waist-to-hip ratio. Among women, 81.7% had a high-risk waist circumference. These findings are consistent with data reported in Morocco by El Rhazi et al., who observed abdominal obesity in approximately 62% of adult women in urban settings, as well as with the findings of Macia et al. in Dakar, where abdominal obesity affected 37.5% of women compared with 5.5% of men [11,12]. These findings are important for cancer prevention, as excess body weight and abdominal adiposity are recognized risk factors for several cancer sites. The IARC Working Group, as reported by Lauby-Secretan et al., confirmed the role of adiposity in several cancers, while the World Cancer Research Fund highlights the association between abdominal adiposity and colorectal, pancreatic, and postmenopausal breast cancers[2,7]. Although the present study does not allow causal relationships to be established, it highlights a high prevalence of modifiable anthropometric risk factors in Nouakchott, particularly among women.

Beyond anthropometric indicators, the dietary profile observed in the present study was characterized by frequent exposure to several unfavorable dietary factors. Red meat consumption was high, with 40.4% of participants reporting daily intake and 66.8% consuming it at least three times per week. In Morocco, available regional data suggest frequent red meat consumption among adults, although direct comparisons remain limited because of differences in exposure measurement [13]. Processed meat consumption was lower in the present study but remained notable, with 15.1% of participants reporting consumption at least three times per week.

These findings are relevant because the World Cancer Research Fund recommends limiting red meat intake and consuming very little, if any, processed meat [7]. The International Agency for Research on Cancer classified processed meat as carcinogenic to humans and red meat as probably carcinogenic to humans, mainly on the basis of evidence related to colorectal cancer [14,15]. Although the present study assessed consumption frequency rather than quantity, the high frequency of red meat consumption in Nouakchott represents a significant exposure warranting preventive action.

The intake of foods considered protective was uneven. Daily fresh fruit consumption was reported by only 13.8% of participants, while 20.9% reported no fruit consumption. Daily fresh vegetable consumption was more frequent, reported by 43.4%, but 23.4% reported no vegetable consumption. These results are consistent with WHO STEPS Mauritania 2006, which reported that 94.8% of adults in Nouakchott consumed fewer than five combined servings of fruit and vegetables per day [16]. They are also comparable with the Morocco STEPS 2017 survey, which reported that 76.3% of adults consumed fewer than five servings of fruit and vegetables per day [17]. These data confirm that insufficient consumption of plant-based foods remains common in North African settings.

Regarding dairy products, the regular consumption observed in the present study is consistent with some regional data. In Tunisia, Soethoudt et al. reported in 2018 an average consumption of approximately 110 liters of dairy products per capita per year, while Zlaoui et al. showed in 2021 that 67% of consumers purchased milk and 72% purchased yogurt two to four times per week. [18,19]. From a prevention perspective, the World Cancer Research Fund reports an association between dairy products, calcium intake, and reduced colorectal cancer risk. Similarly, Keum and Giovannucci indicated that an increase in calcium intake of approximately 300 mg/day was associated with an approximately 6% reduction in colorectal cancer risk [7,20]. These findings should nevertheless be interpreted with caution, as the present study measured consumption frequency rather than quantities consumed. Overall, potentially protective foods were present in the diet of the study population, but unevenly, with a more marked insufficiency in fresh fruit consumption.

Sugar-related exposures were also frequent. Foods rich in refined sugars were consumed daily by 39.9% of participants, and sugar-sweetened beverages were consumed at least weekly by 61.7%, including 14.9% reporting daily consumption. In Africa, Karugu et al. reported in 2025 increasing sales of sugar-sweetened beverages across several African countries and a growing contribution to cardiometabolic risk [21]. In the French NutriNet-Santé cohort, Chazelas et al. reported in 2019 that a 100 mL/day increase in sugar-sweetened beverage consumption was associated with an 18% increase in overall cancer risk [22]. Similarly, Llaha et al., in a systematic review and meta-analysis published in 2021, concluded that higher consumption of sugar-sweetened beverages was associated with an increased risk of several cancers [23].

Overall, the nutritional profile observed in Nouakchott combines high exposure to red meat, refined cereals, sugary foods and sugar-sweetened beverages with insufficient daily fruit intake. This supports nutrition education strategies promoting fruits, vegetables, legumes, fiber-rich foods and minimally processed foods, while reducing excessive consumption of red meat, processed foods and added sugars.

In addition to nutritional exposures, behavioral risk factors were also highly prevalent. Physical inactivity was among the most prevalent behavioral risk factors identified in this study. Overall, 64.7% of participants reported no regular physical activity, and only 4.6% reported physical activity five or more times per week. This level is higher than the WHO STEPS Mauritania 2006 estimate, where low physical activity was reported in 50.7% of adults in Nouakchott [16]. At a broader scale, Guthold et al. showed in 2008, in a survey covering 51 countries, that Mauritania had a high prevalence of physical inactivity, reaching 51.7% among men and 71.2% among women [24]. In Arab countries, Sharara et al. reported in 2018 that the prevalence of physical inactivity among adults often exceeded 40% [25].

This finding has direct implications for cancer prevention. Moore et al., in a large prospective study including 1.44 million adults in 2016, showed that higher leisure-time physical activity was associated with reduced risk of 13 types of cancer, including colorectal, breast, endometrial, liver and esophageal cancers [26]. In Nouakchott, physical inactivity probably interacts with the high prevalence of excess weight and abdominal adiposity, which reinforces the need for integrated health promotion interventions.

Current smoking was reported by 15.7% of participants. This prevalence is slightly lower than that reported in WHO STEPS Mauritania 2006, where 18.9% of adults in Nouakchott were current smokers, including 34.2% of men and 5.7% of women [16].

In the Maghreb context, reported levels vary. In Morocco, the 2017 STEPS survey reported a current smoking prevalence of 11.7% among adults, whereas in Tunisia, the 2016 Tunisian Health Examination Survey reported a prevalence of 22.3% among adults aged 15 years and older. The prevalence observed in the present study therefore falls between the levels reported in Morocco and Tunisia [17,27]. The carcinogenic role of tobacco is well established. The World Health Organization emphasizes that tobacco smoke contains numerous carcinogens and is involved in several cancers [28]. Reitsma et al., in the GBD 2019 analysis, confirmed that smoking remains one of the leading preventable contributors to morbidity and mortality worldwide [29].

In Nouakchott, the persistence of current smoking supports the need to strengthen tobacco control, particularly prevention of initiation, cessation support and awareness-raising.

Daily sun exposure was very frequent, reported by 98.4% of participants, while only 1.6% reported no exposure. The most common duration was less than 30 minutes per day, reported by 39.1%, but 25.4% reported exposure of at least one hour per day. These findings should be interpreted in the Sahelian context of Mauritania, characterized by high sunlight intensity. The World Health Organization states that excessive ultraviolet radiation is a major risk factor for skin cancers, and that more than 1.5 million cases of skin cancer were diagnosed worldwide in 2020 [30]. The carcinogenic effects of ultraviolet radiation are mainly based on DNA damage, oxidative stress, and alterations in cellular repair mechanisms [31].

Although the present study did not assess cumulative lifetime exposure or ultraviolet intensity, the high frequency of sun exposure supports prevention messages on sun protection for highly exposed groups.

The stratified analysis by sex, age group and wilaya of residence provided additional insights into the distribution of these risk factors. Sex differences were mainly anthropometric. Women were more frequently obese than men, 40.0% versus 15.0%, and more frequently had high-risk waist circumference, 81.7% versus 32.3%. Conversely, elevated waist-to-hip ratio was more common among men, 67.5% versus 36.5%. This pattern is consistent with regional evidence showing a greater burden of obesity and abdominal adiposity among women in several African and Maghreb settings [9,12]. In contrast, no significant differences by sex were observed for the main high-consumption dietary variables, suggesting that sex differences were more related to body size and fat distribution than to declared dietary frequency.

Age-related differences mainly concerned selected dietary habits. High red meat consumption increased from 51.4% among participants aged 18–24 years to 75.6% among those aged 55–64 years. High fresh vegetable consumption was highest among participants aged 45–54 years, at 62.9%, while high dairy product consumption was highest among those aged 65 years and older, at 67.7%. However, body mass index categories and regular physical activity did not vary significantly by age. These findings suggest that exposure to several nutritional risk factors is already present among young adults, supporting prevention across the life course.

Differences by wilaya mainly concerned behavioral and dietary factors. Regular physical activity was lowest in Nouakchott West, at 24.0%, while current smoking was highest in Nouakchott South, at 18.6%. High processed meat and canned food consumption were more frequent in Nouakchott North, at 19.0% and 30.9%, respectively, whereas high fresh vegetable consumption was highest in Nouakchott South, at 57.2%. These intra-urban differences indicate that prevention interventions should be tailored to the specific exposure profiles of each wilaya rather than applied uniformly.

This study has several strengths that merit consideration. It is one of the first studies conducted in Mauritania on nutritional, behavioral and anthropometric cancer-related risk factors in the general adult population. It included 1,000 adults from the three wilayas of Nouakchott and assessed several complementary dimensions of risk, including diet, physical activity, smoking, sun exposure and anthropometric indicators.

Some limitations should be acknowledged. The cross-sectional design does not allow causal relationships to be established. Several variables relied on self-report, which may have introduced recall bias and/or social desirability bias. Dietary habits were measured as frequency of consumption rather than quantities consumed, limiting comparison with studies based on grams, portions or nutrient intake. Finally, the high proportion of participants with higher education (61.2%) may reflect a selection bias inherent to urban sampling in Nouakchott, where educational institutions and administrative employment are concentrated, as well as the use of a digital data collection tool (KoboCollect) that may have favored more educated and digitally literate respondents. This limits the generalizability of findings to less educated or rural Mauritanian populations and may lead to an underestimation of some risk factors more prevalent in lower-education groups.

From a public health perspective, these findings highlight the need to strengthen primary prevention of cancer and other noncommunicable diseases in Nouakchott. Priority actions should include prevention of excess body weight, promotion of regular physical activity, reduction of smoking, limitation of red meat, processed foods and sugar-sweetened beverages, and promotion of fruit, vegetables, legumes and fiber-rich foods. The observed differences by sex, age and wilaya indicate that prevention should be targeted rather than uniform, with particular attention to obesity and abdominal adiposity among women and to local exposure patterns in each wilaya.

## 5 Conclusion

The present cross-sectional study conducted among 1,000 adults in the three wilayas of Nouakchott highlights a high prevalence of several nutritional and behavioral cancer-related risk factors in the general population. The most notable findings concern the high prevalence of excess body weight, abdominal adiposity, physical inactivity, and selected unhealthy dietary habits, particularly frequent consumption of red meat, sugar-sweetened beverages, and refined cereals. Conversely, the consumption of potentially protective foods appeared uneven, with a more marked insufficiency in fresh fruit consumption.

These findings reflect a concerning exposure profile in an urban context undergoing nutritional and epidemiological transition. They indicate that cancer prevention in Nouakchott should be integrated into a broader approach to controlling noncommunicable diseases, focused on modifiable risk factors. Strengthening primary prevention through the promotion of a more balanced diet, prevention of excess body weight, encouragement of regular physical activity, and reduction of smoking should be considered a priority.

Finally, this study provides useful local data to guide public health interventions. Further research, ideally multicenter and longitudinal, is needed to monitor trends in these risk factors, better identify their determinants, and guide national policies for the prevention of cancer and other chronic diseases.

These findings may apply mainly to adults in urban settings undergoing nutritional transition and may not be directly generalizable to rural Mauritanian populations.

## Data Availability

The anonymized dataset supporting the findings of this study is publicly available on Zenodo at: https://doi.org/10.5281/zenodo.20289078

## 6 Declarations

### Ethics approval and consent to participate

This study was conducted in accordance with the ethical principles of the Declaration of Helsinki. The study protocol was approved by the Health Research Ethics Committee of the Ministry of Health of Mauritania under reference number 047-2026/MS/CNERS, dated January 22, 2026. Informed consent was obtained from all participants before inclusion in the study.

### Consent for publication

Not applicable.

### Authors’ contributions

NT contributed to study conception, study design, field coordination, data analysis, interpretation of results and drafting of the manuscript. All co-authors contributed to scientific supervision, interpretation of findings and critical revision of the manuscript. All authors read and approved the final manuscript.

## Acknowledgements

The authors would like to thank all study participants for their cooperation. The authors also thank the data collection team and the health authorities who facilitated the implementation of the survey in the three wilayas of Nouakchott.

## Use of artificial intelligence tools

An artificial intelligence-assisted tool was used only to support translation of the manuscript into English and language editing. It was not used for study design, data collection, data cleaning, statistical analysis, interpretation of results, formulation of conclusions, or generation of scientific content. All scientific content, analyses and interpretations were developed and verified by the authors. All authors reviewed, edited and approved the final manuscript and take full responsibility for its content.

